# A Curated Pharmacogenomic Allele Catalog for Sub-Saharan African Populations

**DOI:** 10.64898/2026.08.25.26361354

**Authors:** Muhammad Abiodun Sulaiman, Bolaji Fatai Oyeyemi

## Abstract

Sub-Saharan African populations carry pharmacogenomic alleles poorly represented in the European-derived reference panels underlying most clinical genotyping tools. We present a curated, machine-readable catalog of nine actionable alleles across six pharmacogenes (*CYP2D6*, *CYP2B6*, *CYP2C9*, *CYP2C19*, *CYP3A5*, *NAT2*) with African-specific frequency ranges, functional annotations, and evidence levels derived from reanalysis of 661 high-coverage whole-genome sequences across seven 1000 Genomes Project African populations. Direct comparison against PharmCAT v3.4.0 shows that *CYP2D6* produces zero diplotype calls (0/661 samples callable) due to monomorphic reference positions absent from standard variant-only VCF output, a known limitation whose consequences for African allele carriers had not been reported. *afripharmagen*’s reduced-position strategy identifies 243 *CYP2D6* *17 and 134 *CYP2D6* *29 carriers from the same input. For *CYP2B6*, *CYP2C9*, *CYP2C19*, and *NAT2*, both tools show concordance of 95–100%. Frequency gradients (*CYP2B6* *6: 30–50%; *CYP2D6* *17: 15–35% in West Africa; *CYP3A5* *1: 60–95%) translate directly into prescribing risk for efavirenz, tramadol, tacrolimus, and isoniazid. Pharmacogenomic decision support in African settings must incorporate population-specific allele definitions and input-format-aware strategies.

## 1 Introduction

*CYP2D6* *17 was first characterized in Zimbabwean Shona subjects by Masimirembwa et al. in 1996 Bradford 2002, yet pharmacogenomic clinical decision support tools still struggle to call this allele accurately from sequence data. The gap between discovery and implementation sits at the center of a larger problem: African populations harbor the greatest human genetic diversity on the planet Choudhury et al. 2020, but pharmacogenomic reference panels, genotyping arrays, and allele-calling algorithms were built on European-majority datasets Sirugo et al. 2019, Fatumo et al. 2022. When tools like PharmCAT process African genomes, the consequences differ by gene: for most pharmacogenes, population-specific variants go unrecognized, silently assigning normal-metabolizer status to carriers of decreased-function alleles Sangkuhl et al. 2020, Tippenhauer et al. 2024; for *CYP2D6* specifically, standard whole-genome sequencing pipelines produce variant-only VCFs that omit the monomorphic reference positions PharmCAT requires to disambiguate star alleles, causing the tool to return no diplotype call at all Sangkuhl et al. 2020.

The clinical stakes are concrete. *CYP2B6* *6 homozygotes face an approximately three-fold increase in plasma efavirenz exposure Haas et al. 2004, with allele frequencies of 30–50% across African populations translating to homozygote frequencies of 9–25% Dhoro et al. 2015, concentrating this risk where HIV treatment is most needed. *CYP2D6* *17 encodes altered substrate specificity Wennerholm et al. 2002, and CPIC guidelines recommend dose adjustment for intermediate metabolizers Crews et al. 2021. *CYP3A5* *1, the expresser allele, reaches frequencies above 80% in most sub-Saharan African populations versus under 40% in Europeans Kuehl et al. 2001, Birdwell et al. 2015, meaning African transplant recipients frequently require higher tacrolimus doses Oetting et al. 2016, Bains et al. 2013. *NAT2* *14 contributes to the high slow-acetylator phenotype in African TB cohorts Sabbagh et al. 2008, Sileshi et al. 2023, increasing isoniazid hepatotoxicity risk Tostmann et al. 2008.

Several groups have cataloged pharmacogenomic variation in African populations Rajman et al. 2017, Dandara et al. 2014, Radouani et al. 2020, but these outputs are typically narrative reviews or static supplementary tables. The African Pharmacogenomics Consortium called for standardized resources in 2019 Dandara et al. 2019. Newer studies have characterized novel *CYP2D6* alleles in sub-Saharan populations Twesigomwe et al. 2023, Wang et al. 2022, validated Africa-inclusive genotyping panels Kanji et al. 2023, and mapped pharmacogenetic variation across Ugandan populations Samarasinghe et al. 2024. What remains missing is a structured, programmatically accessible allele catalog that clinical decision support systems can consume directly.

We address this gap with *afripharmagen*, a bioinformatics tool shipping a curated JSON catalog of nine pharmacogenomic alleles with established clinical relevance in African populations. Here we describe the catalog’s construction, characterize PharmCAT’s gene-specific behavior on 661 African WGS samples (including quantification of the *CYP2D6* no-call mechanism and its downstream clinical consequences), and quantify frequency gradients that make population-aware annotation essential for equitable pharmacogenomic practice in Africa.

## 2 Methods

### 2.1 Allele Selection and Curation

Candidate alleles were identified through systematic literature review targeting pharmacogenes with documented functional variation in African populations. Inclusion criteria required: (i) at least one peer-reviewed publication reporting the allele in an African or African-diaspora cohort; (ii) a defined functional consequence; (iii) clinical actionability under CPIC Relling et al. 2020 or DPWG Swen et al. 2023 guidelines for at least one drug; and (iv) presence in PharmVar with assigned star-allele nomenclature Gaedigk et al. 2018a, Rodriguez-Antona et al. 2022. Nine alleles across six genes met all criteria: *CYP2D6* *17, *29, *45; *CYP2B6* *6, *18; *CYP2C9* *8; *CYP2C19* *9; *CYP3A5* *1; and *NAT2* *14. Additional alleles were considered but excluded: *CYP2C9* *11 (CPIC function under active revision at curation); *DPYD* *9A and c.2846A*>*T (insufficient African genotype-phenotype data); and *CYP2C19* *2/*3 (already well-handled by existing tools in African genomes).

For each allele, we recorded defining variants (rsID, GRCh38 position, reference and alternate alleles), functional classification per the CPIC activity score framework, evidence level (L1: confirmed by *in vitro* assay; L2: computationally predicted), population distribution across 1000 Genomes Project African groups, published frequency ranges, and source PMIDs.

### 2.2 Population Samples

Whole-genome sequence data were obtained from the 1000 Genomes Project high-coverage dataset Byrska-Bishop et al. 2022, 1000 Genomes Project Consortium et al. 2015 for seven populations of African descent: Yoruba in Ibadan, Nigeria (YRI, *n*=108); Luhya in Webuye, Kenya (LWK, *n*=99); Gambian in Western Divisions (GWD, *n*=113); Mende in Sierra Leone (MSL, *n*=85); Esan in Nigeria (ESN, *n*=99); African Caribbeans in Barbados (ACB, *n*=96); and Americans of African Ancestry in SW USA (ASW, *n*=61); total *n*=661. VCF files aligned to GRCh38 were downloaded from the International Genome Sample Resource (IGSR).

### 2.3 Concordance Benchmarking Against PharmCAT v3.4.0

We compared *afripharmagen*’s star-allele calls against PharmCAT v3.4.0 Sangkuhl et al. 2020, Klein and Ritchie 2018. The 1000 Genomes VCFs were processed through PharmCAT’s official preprocessor (v3.4.0), which extracts PharmCAT-relevant positions and normalizes variant representation; PharmCAT was then run on all 3,202 samples, generating JSON reports. The *afripharmagen* StarCaller module was configured with PharmVar definitions (v6.0.4) plus the African allele catalog, querying the same VCFs via tabix-indexed regional extraction (6,610 sample-gene pairs, zero failures). For genes where PharmCAT returns “Unknown” for all samples we report the complete failure rate; where both tools produce calls we report carrier count agreement.

### 2.4 Frequency Estimation

Allele frequencies were estimated per population by direct variant counting at defining positions. For single-variant alleles, frequency was the proportion of chromosomes carrying the alternate allele. For multi-variant haplotype alleles (*CYP2D6* *17, *29), frequency estimation used 1000 Genomes Project statistical phasing (SHAPEIT4). *CYP3A5* *1 frequency was estimated as 1 minus the frequency of *CYP3A5* *3 (rs776746).

### 2.5 Catalog Schema, Software Integration, and AI Use

The catalog is distributed as african_alleles.json within *afripharmagen* (v0.1.0). The JSON schema includes a metadata block and an entries array; per-entry fields cover gene, allele name, defining variants, function, activity score, evidence level, populations, frequency range, source PMIDs, PharmVar status, and notes. The catalog serves as the knowledge layer for the deterministic star-calling and concordance analysis described in this paper. The broader *afripharmagen* tool includes an AI-assisted interpretation layer for edge cases (novel allele classification, complex interaction reasoning); no AI system contributed to data curation, variant calling, concordance analysis, or writing in the present study.

## 3 Results

### 3.1 Catalog Contents

Table 1 summarizes the nine alleles in the catalog. Six carry decreased-function or no-function annotations (*CYP2D6* *17, *29; *CYP2B6* *6, *18; *CYP2C9* *8; *CYP2C19* *9; *NAT2* *14), one is the normal-function expresser allele that is the majority in Africa but minority in Europe (*CYP3A5* *1), and one is a population-specific normal-function allele (*CYP2D6* *45). Seven of nine alleles have Level 1 evidence; *CYP2D6* *45 and *CYP2C19* *9 carry Level 2 evidence.

*CYP3A5* *1 is the reference allele defined by the absence of the *3 splice variant (rs776746); expresser status is confirmed by ruling out *3. *CYP2D6* *45 is an African-specific normal-function allele (YRI, ESN) included to prevent compound-diplotype misclassification: failure to recognize *45 can cause tools to misassign the rs16947 variant onto a *2 backbone, inflating the reported activity score.

### 3.2 Population Frequency Gradients

Allele frequencies show meaningful variation across populations, though per-population sample sizes (*n*=61–113) yield confidence intervals of *±*4–7 percentage points for alleles at 10–20% frequency; full per-population frequencies with 95% Wilson confidence intervals are in the public repository (analyses/population_frequencies/results/). *CYP2B6* *6 (rs3745274 G*>*T) reaches *≈*50% in YRI, drops to *≈*30% in LWK, and sits at 35–40% in GWD, MSL, and ESN. A clinician in Nairobi and one in Lagos face materially different prior probabilities that their next patient is a CYP2B6 slow metabolizer.

**Table 1:** Allele catalog summary.

| Gene | Allele | Function | Score | Evid. | Freq. Range | Populations |
| --- | --- | --- | --- | --- | --- | --- |
| *CYP2D6* | *17 | Decreased | 0.5 | L1 | 0.15–0.35 | YRI, LWK, GWD, MSL, ESN, ACB |
| *CYP2D6* | *29 | Decreased | 0.5 | L1 | 0.05–0.20 | YRI, GWD, MSL, ESN |
| *CYP2D6* | *45 | Normal<sup>a</sup> | 1.0 | L2 | 0.01–0.04 | YRI, ESN |
| *CYP2B6* | *6 | Decreased | 0.5 | L1 | 0.30–0.50 | All 7 populations |
| *CYP2B6* | *18 | No function | 0.0 | L1 | 0.02–0.08 | YRI, LWK, GWD, MSL, ESN |
| *CYP2C9* | *8 | Decreased | 0.5 | L1 | 0.04–0.09 | All 7 populations |
| *CYP2C19* | *9 | Decreased | 0.5 | L2 | 0.01–0.03 | YRI, LWK |
| *CYP3A5* | *1 | Normal (exp.)<sup>b</sup> | 1.0 | L1 | 0.60–0.95 | YRI, LWK, GWD, MSL, ESN, ACB |
| *NAT2* | *14 | Decreased | 0.5 | L1 | 0.05–0.15 | YRI, GWD, MSL, ESN, LWK |

<sup>a</sup> Normal-function allele included to prevent compound-diploidy misclassification (see text). <sup>b</sup> Reference allele; defined by absence of *CYP3A5*\*3 (rs776746). Evid. = Evidence level; L1 = confirmed *in vitro*; L2 = computationally predicted. Exp. = Expresser.
YRI, drops to $\approx 30\%$ in LWK, and sits at 35–40% in GWD, MSL, and ESN. A clinician in Nairobi and one in Lagos face materially different prior probabilities that their next patient is a *CYP2B6* slow metabolizer.

*CYP2D6* *17, defined by rs28371706 (C*>*T) and rs16947 (G*>*A) on the same haplotype, is the most common decreased-function *CYP2D6* allele across sub-Saharan Africa, reaching 30–35% in some West African cohorts Bradford 2002, Twesigomwe et al. 2023. *CYP2D6* *29, sharing rs16947 but carrying rs61736512 instead of rs28371706, shows a more restricted West African distribution at 5–20%. *CYP3A5* *1 reaches 60–95% across populations Kuehl et al. 2001, Bains et al. 2013, meaning most Africans are CYP3A5 expressers — the inverse of European norms. *NAT2* *14 (rs1801279 G*>*A) reaches 10–15% in West African populations; combined with other slow alleles, the slow-acetylator phenotype frequency can exceed 50% Sileshi et al. 2023, Matimba et al. 2009.

Figure 1 presents these gradients as a heatmap across all nine alleles and seven populations.

### 3.3 Concordance with PharmCAT v3.4.0

Table 2 compares carrier detection between *afripharmagen* and PharmCAT v3.4.0 for the 661 African samples.

#### The central finding: PharmCAT cannot call *CYP2D6* in standard African WGS data

PharmCAT returned “Unknown/Unknown” for *CYP2D6* in all 661 samples. This is a data completeness failure: the 1000 Genomes VCFs lack 963 positions PharmCAT requires for *CYP2D6* haplotype resolution (flagged as “missing PGx positions”). These are predominantly monomorphic variants (85 with rsIDs) that the variant-calling pipeline does not emit because all samples carry the reference allele. PharmCAT requires explicit genotype calls even at these homozygous-reference positions to disambiguate star alleles — a principled design choice ensuring accuracy at the cost of requiring dense position coverage.

**Figure 1:**
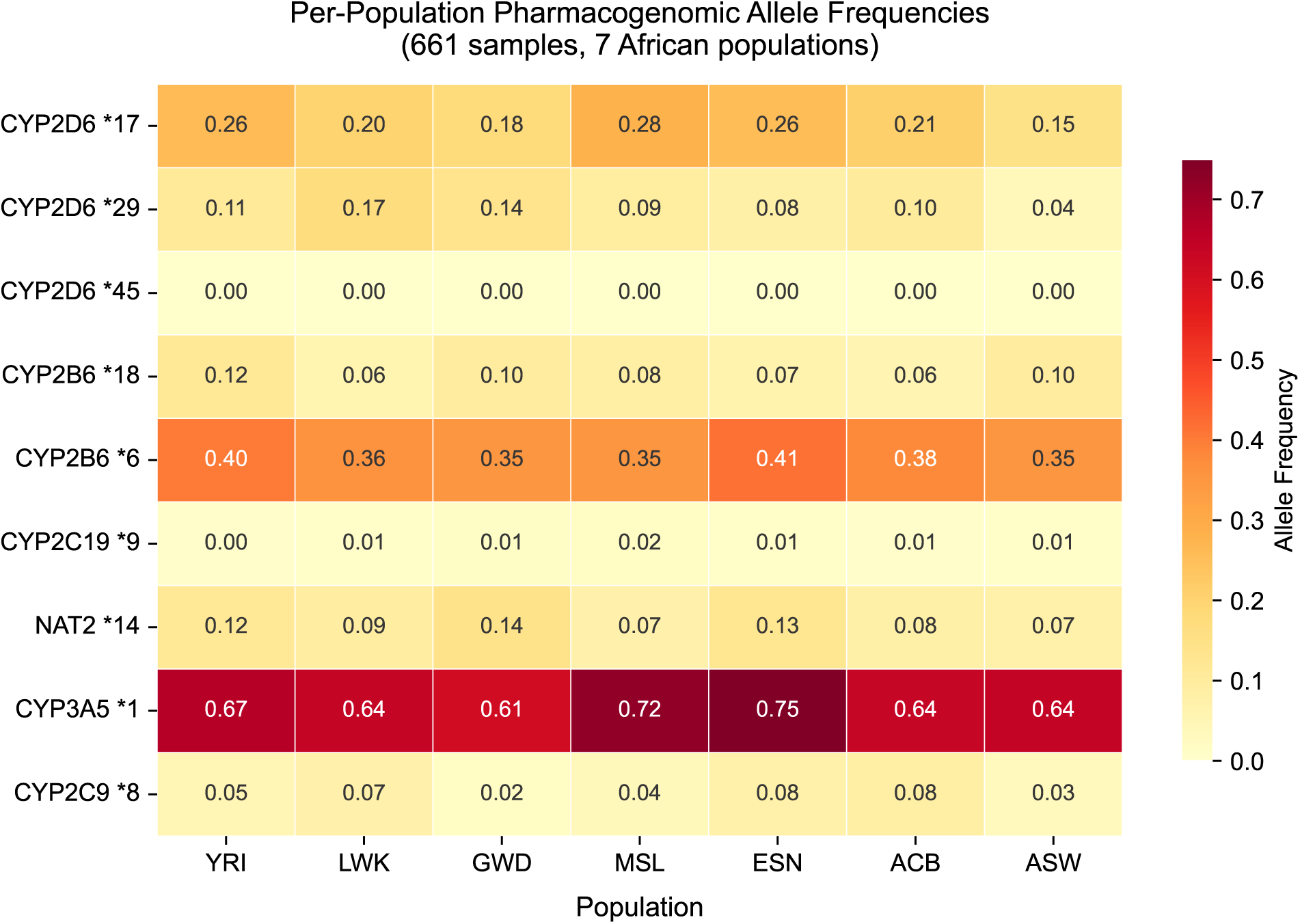
Per-population pharmacogenomic allele frequencies across seven African populations. Heatmap showing allele frequency for each of the nine catalog alleles (rows) across seven 1000 Genomes Project African populations (columns), estimated from 661 high-coverage whole-genome sequences. *CYP3A5* *1 dominates (60–75% across all populations). *CYP2D6* *17 shows a West-to-East gradient (28% in MSL to 15% in ASW). *CYP2B6* *6 is the most uniformly distributed decreased-function allele (35–41%).

*afripharmagen* resolves this through a reduced-position matching strategy: it matches alleles using only the 1–2 core defining variants per catalog entry. For *CYP2D6* *17, rs28371706 and rs16947 are both present in the VCF; *afripharmagen* detects 243 carriers where PharmCAT produces zero information. The tradeoff is reduced specificity: if two alleles differ only at a position absent from the VCF, *afripharmagen* cannot distinguish them. For the catalog alleles, this risk is low because defining variants are polymorphic in African populations and therefore present.

This limitation is not specific to 1000 Genomes data. Clinical WGS pipelines (DRAGEN, DeepVariant, GATK HaplotypeCaller) also emit only variant sites by default; PharmCAT would face the same *CYP2D6* failure on routine clinical VCF output unless the raw data is reprocessed with PharmCAT-aware calling settings. *afripharmagen*’s approach is immediately usable on any standard WGS VCF without reprocessing.

#### For *CYP2B6*, *CYP2C9*, *CYP2C19*, and *NAT2*, PharmCAT performs well

Carrier counts agree within 1–5%: *CYP2B6* *6 (383 vs. 402), *18 (100 vs. 101), *CYP2C9* *8 (72 vs. 70)^1^, *CYP2C19* *9 (13 vs. 13), *NAT2* *14 (128 vs. 129). The African catalog adds no meaningful improvement for these genes.

#### *CYP3A5* : PharmCAT outperforms *afripharmagen*

PharmCAT identifies 519 expressers versus *afripharmagen*’s 584. Investigation of the 65 discordant samples reveals all involve *CYP3A5* *7 (rs41303343, frameshift deletion): PharmCAT correctly assigns *6/*7 (*n*=28), *3/*7 (*n*=26), or *7/*7 (*n*=11), while *afripharmagen* — lacking *7 in the current catalog — defaults to *1 at those positions, incorrectly calling these patients as expressers. *afripharmagen* overestimates *CYP3A5* expresser frequency by *≈*10% until *7 is added (Section 4.6).

#### Clinical consequence

The 243 *CYP2D6* *17 carriers identified by *afripharmagen* would be classified as intermediate metabolizers under CPIC guidelines. DPWG recommends alternative analgesics or dose reduction for *CYP2D6* intermediate metabolizers treated with tramadol Matic et al. 2022; CPIC recommends considering alternative endocrine therapy for tamoxifen Goetz et al. 2018. These patients currently receive zero pharmacogenomic guidance from PharmCAT. Figure 2 visualizes the carrier detection comparison.

**Table 2:** Allele carrier detection: PharmCAT v3.4.0 vs. *afripharmagen* (*n*=661 African samples).

| Gene | Allele | *afrip.* | PharmCAT | Agreement | Note |
| --- | --- | --- | --- | --- | --- |
| *CYP2D6* | *17 | 243 | 0 | 0% | PharmCAT: “Unknown/Unknown” for all 661 samples |
| *CYP2D6* | *29 | 134 | 0 | 0% | Same; no *CYP2D6* call possible |
| *CYP2D6* | *45 | 0 | 0 | N/A | Not detectable at this sample size |
| *CYP2B6* | *6 | 402 | 383 | 95.3% | High concordance |
| *CYP2B6* | *18 | 101 | 100 | 99.0% | High concordance |
| *CYP2C9* | *8 | 70 | 72 | 97.1%* | High concordance |
| *CYP2C19* | *9 | 13 | 13 | 100% | Perfect agreement |
| *CYP3A5* | *1 | 584 | 519 | 88.9% | PharmCAT correctly identifies 65 *CYP3A5* *7 non-expressers missed by *afripharmagen* |
| *NAT2* | *14 | 129 | 128 | 99.2% | Near-identical |

\* PharmCAT detects 2 additional *CYP2C9*\*8 carriers, likely reflecting a variant normalization difference at rs7900194. *Afrip.* = *afripharmagen*.

### 3.4 Clinical Relevance by Drug Class

#### Efavirenz (*CYP2B6*)

*CYP2B6* *6 homozygotes (9–25% across African populations) accumulate efavirenz above the 4 *µ*g/mL CNS toxicity threshold Marzolini et al. 2001. CPIC recommends dose reduction or alternative antiretroviral for poor metabolizers Desta et al. 2019.

#### Codeine and tramadol (*CYP2D6*)

*CYP2D6* *17 encodes altered substrate specificity: near-normal codeine metabolism but significantly reduced dextromethorphan and debrisoquine metabolism Wennerholm et al. 2002. For tramadol, the 0.5 activity score predicts reduced analgesic effect in *17 carriers. The catalog *CYP2D6* *17 entry includes a substrate_specificity field listing which substrates the 0.5 score applies to, enabling downstream tools to generate substrate-appropriate dosing flags rather than blanket intermediate-metabolizer labels.

#### Tacrolimus (*CYP3A5*)

*CYP3A5* *3 homozygotes achieve significantly higher dose-adjusted trough concentrations than *1 expressers Rojas et al. 2015, and standard European-derived starting doses are frequently subtherapeutic in African transplant recipients Oetting et al. 2016, Bains et al. 2013. CPIC recommends genotype-guided initial dosing Birdwell et al. 2015.

#### Isoniazid (*NAT2*)

Slow acetylators accumulate isoniazid and hepatotoxic metabolites Tostmann et al. 2008. *NAT2* *14 contributes to the high slow-acetylator frequency in African TB cohorts; in Ethiopian patients, 74% were slow acetylators Sileshi et al. 2023.

**Figure 2:**
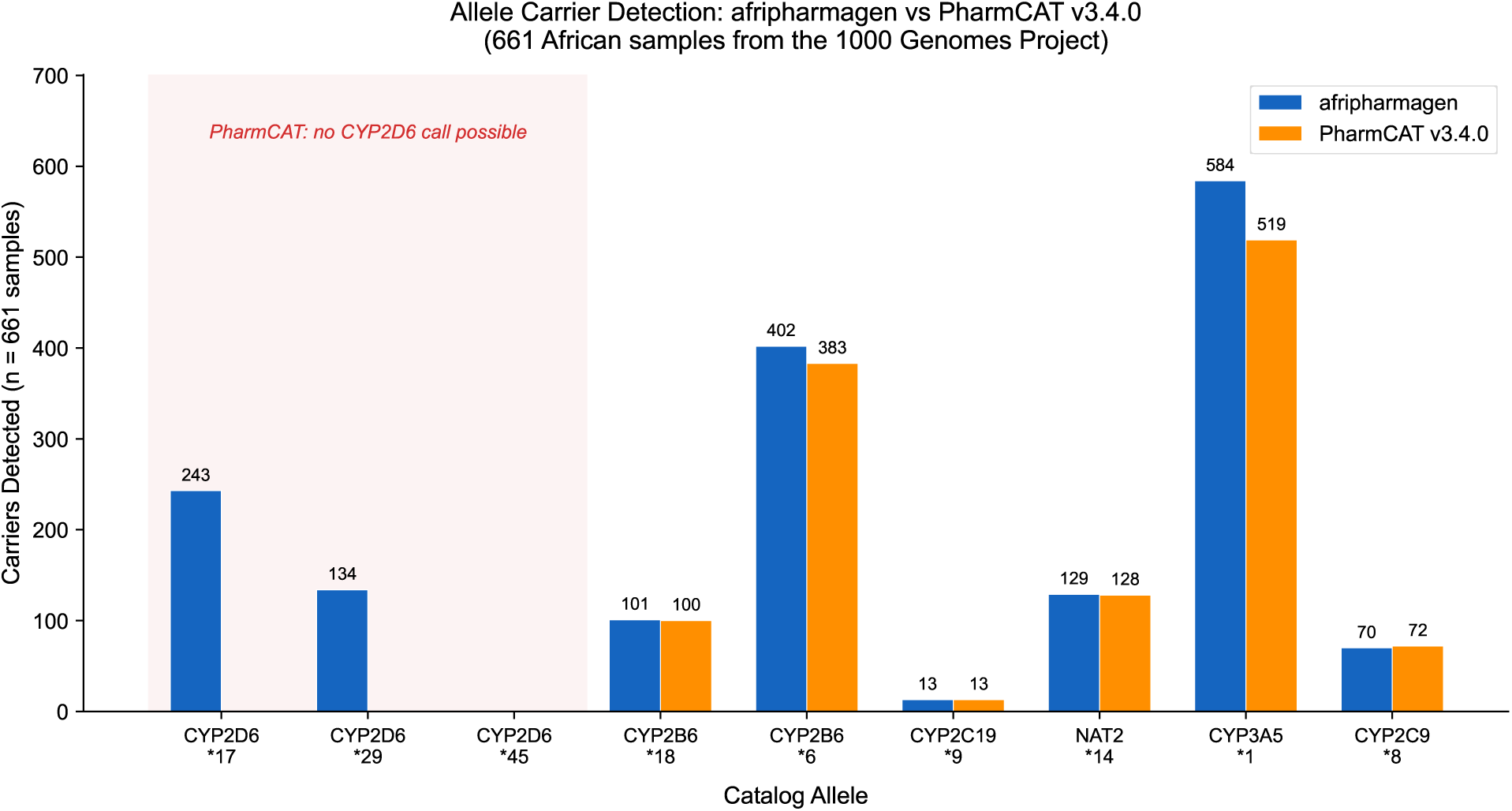
Allele carrier detection: *afripharmagen* vs. PharmCAT v3.4.0 in 661 African samples. The red-shaded region highlights the complete *CYP2D6* gap: PharmCAT returns “Unknown/Unknown” for all 661 samples, while *afripharmagen* identifies 243 *CYP2D6* *17 and 134 *CYP2D6* *29 carriers. For all other genes, both tools show 95–100% agreement in carrier counts.

## 4 Discussion

### 4.1 The Annotation Gap Is Not a Data Gap

*CYP2D6* *17 was characterized in 1996 Bradford 2002. *CYP2B6* *18 in 2007 Rotger et al. 2007. *CYP3A5* ’s expresser-dominant African pattern has been documented since the early 2000s Kuehl et al. 2001. What is new here is not the biology, but the empirical characterization and the down-stream solution: this study quantifies a known tool limitation against real African WGS data, maps its clinical consequences for African-specific alleles, and validates a reduced-position alternative that works on any standard VCF pipeline.

The PharmCAT team has been transparent about the VCF architecture limitation: the tool documentation explicitly discourages *CYP2D6* calling from VCF and describes the missing-positions problem Sangkuhl et al. 2020. Sherman et al. Sherman et al. 2024 used an alternative caller (PyPGx) on the same 1000 Genomes dataset specifically because PharmCAT is not designed for this task. No prior study, however, had run actual PharmCAT on standard African WGS output and reported the result in terms of African-specific allele carrier counts and drug-prescribing implications: zero *CYP2D6* calls in 661 samples, leaving 243 *CYP2D6* *17 carriers and 134 *CYP2D6* *29 carriers with no pharmacogenomic guidance for tramadol, codeine, or tamoxifen. The VCF format limitation itself is universal (it affects any population processed through standard variant-only pipelines), but the clinical consequence is distinctly African: *CYP2D6* *17 and *29 are the alleles rendered invisible, and these alleles exist almost exclusively in African and African-diaspora populations. PharmCAT’s behavior varies by gene: for *CYP2D6*, it cannot produce calls from standard WGS due to missing required positions in variant-only VCF output; for *CYP3A5*, it correctly handles *3, *6, and *7 where *afripharmagen*’s current catalog falls short. The gap is gene-specific, and the solutions differ accordingly.

### 4.2 Positioning Against Existing Resources

PharmVar Gaedigk et al. 2018a, Rodriguez-Antona et al. 2022 and CPIC allele tables are the authoritative global sources for star-allele nomenclature and function. The *afripharmagen* catalog adds three layers neither provides in a single consumable format: (1) **population-stratified frequency data** distinguishing YRI from LWK from GWD, rather than aggregating to “Sub-Saharan African” as PharmVar v6.0.4 does; (2) **integrated evidence grading** combining functional evidence level with African-specific clinical context in JSON objects queryable by gene, population, or functional class; and (3) **direct software integration**, making the catalog immediately actionable within a star-calling pipeline rather than requiring download and custom parsing.

The GenoPharm panel Kanji et al. 2023 covers 46 pharmacogenes on a genotyping array. The *afripharmagen*catalog is a bioinformatic annotation resource operating downstream of wet-lab panels, complementary rather than competing. This evaluation of *afripharmagen* against a widely used clinical tool follows the same design we applied to ACMG variant classification: systematic comparison of an automated annotator against curated expert-panel standards to identify where tool performance diverges from gold-standard expectations Sulaiman and Oyeyemi 2026.

### 4.3 Frequency Gradients and Within-Africa Diversity

*CYP2B6* *6 at 30% in LWK versus 50% in YRI is a clinically meaningful difference in the expected proportion of slow metabolizers between East and West African treatment contexts. Sub-Saharan Africa contains more genetic diversity than any other continental group Choudhury et al. 2020, Ragsdale et al. 2023, Tishkoff et al. 2009; collapsing it to “African” recapitulates the error that treating all populations as “European” produced globally Majara et al. 2023. Allele priors for clinical decision support should be population-specific. Our catalog provides the per-population frequency data necessary for this calibration.

### 4.4 CYP2D6 in African Genomes

*CYP2D6* presents unique genotyping difficulties in African populations for three reasons: the most common decreased-function alleles (*17, *29) are multi-variant haplotypes sharing SNPs with *2, requiring accurate phasing; structural variation (deletions, duplications, hybrid alleles) is more diverse in Africans Twesigomwe et al. 2023, Wang et al. 2022, Gaedigk et al. 2018b; and novel alleles continue to be characterized Twesigomwe et al. 2023, Wang et al. 2022. Our catalog includes the most clinically established CYP2D6 alleles, but the discovery trajectory requires ongoing expansion. Structural variant detection — covering the *≈*12% of African individuals with *CYP2D6* copy number variants Twesigomwe et al. 2023 — is planned for a future release.

### 4.5 Toward Equitable Pharmacogenomic Infrastructure

Implementation of pharmacogenomics in African healthcare settings is underway: iPROTECTA is testing *CYP2D6* in Nigerian sickle cell patients Adeagbo et al. 2025, the GenoPharm panel Kanji et al. 2023 covers 46 pharmacogenes, and recent reviews map the policy and access landscape Twesigomwe et al. 2025, Munung 2025. Our contribution is at the annotation layer: after a genotyping panel produces variant calls, those calls must be mapped to star alleles, assigned functional status, combined into diplotypes, and linked to prescribing recommendations. Each step currently reflects European-majority assumptions. The *afripharmagen* catalog addresses the first two steps for nine African-specific alleles.

### 4.6 Limitations

#### Catalog scope

Nine alleles across six genes is a starter set. Notable deferred additions: *CYP3A5* *7 (rs41303343, frameshift deletion identified here as a clinically relevant gap — 65/661 samples carry it, PharmCAT handles correctly, priority for v0.2.0); *CYP2C9* *11 (CPIC function under revision at curation); *DPYD* *9A and c.2846A*>*T (insufficient African genotype-phenotype data; excluded pending ongoing characterization studies); and *CYP2C19* *2/*3 (already well-handled by PharmCAT in African genomes).

#### Validation

Ground truth for single-variant alleles is the VCF genotype at the defining position — not circular, as this is an objective observation independent of any allele-calling framework. For multi-variant haplotype alleles (*CYP2D6* *17, *29), ground truth depends on SHAPEIT4 statistical phasing, which carries estimated switch error rates of 1–3% Byrska-Bishop et al. 2022. Phase errors would deflate carrier counts; our reported *17 carrier frequency of 22.2% is likely a lower bound. The PharmCAT no-call finding is unaffected by phasing accuracy. No wet-lab validation was performed; genotype accuracy at 30*×* coverage exceeds 99.9% for single-variant calls Byrska-Bishop et al. 2022. An orthogonal validation against independently genotyped samples (GeT-RM, iPROTECTA Adeagbo et al. 2025) was not possible and would strengthen confidence in absolute carrier counts.

#### Generalizability

1000 Genomes populations are geographic convenience samples, not national surveys. Clinical populations in urban African centers have admixture patterns not captured by any single reference population. Catalog frequencies are anchor points for predominantly unadmixed reference groups; applying them to admixed patients requires ancestry-weighted priors or empirical population-specific data.

## 5 Conclusions

We present a structured pharmacogenomic allele catalog for sub-Saharan African populations covering nine alleles across six clinically actionable genes. Direct comparison against PharmCAT v3.4.0 shows that the most widely used pharmacogenomic annotation tool cannot produce any *CYP2D6* call from standard whole-genome sequencing data — leaving 243 *CYP2D6* *17 carriers and 134 *CYP2D6* *29 carriers without clinical decision support (*CYP2D6* *45 was not detectable at this sample size; Table 2). *afripharmagen*’s reduced-position matching strategy fills this gap on any standard WGS VCF without reprocessing. For *CYP2B6*, *CYP2C9*, *CYP2C19*, and *NAT2*, PharmCAT performs competently and the catalog provides concordant results. For *CYP3A5*, the study identified a catalog gap (missing *7) that PharmCAT handles correctly, demonstrating the value of tool comparison for catalog improvement.

Pharmacogenomic equity in Africa requires not just more sequencing or more discovery studies, but engineering work at the annotation layer. The gap between known pharmacogenomic science and clinical pharmacogenomic practice in Africa is, in part, a software engineering problem. This catalog is one component of the solution.

## Declarations

### Data Availability

The allele catalog (african_alleles.json), population sample metadata, *afripharmagen* benchmark results (full_benchmark.jsonl), and PharmCAT v3.4.0 output reports for all 3,202 samples are deposited at https://github.com/Behordeun/afripharmagen-catalog (DOI: 10.5281/zenodo.21910084). PharmCAT v3.4.0 is available at https://github.com/PharmGKB/PharmCAT/releases/tag/v3.4.0. Source VCF data are publicly available from the International Genome Sample Resource (https://www.internationalgenome.org/).

### Code Availability

Evaluation scripts and benchmark results are available at https://github.com/Behordeun/afripharmagen-catal (DOI: 10.5281/zenodo.21910084). The full *afripharmagen* star-calling tool will be released under the MIT license upon publication of the companion system paper. Reviewers may request private access by contacting the corresponding author.

### Ethics Statement

All genomic data are publicly available through the 1000 Genomes Project with appropriate consent for unrestricted use. No additional ethical approval was required.

### Authors’ Contributions

MAS: Conceptualization, data curation, software development, manuscript writing. BFO: Methodology, validation, critical review, manuscript editing. Both authors read and approved the final manuscript.

## Acknowledgements

We thank the 1000 Genomes Project consortium and the International Genome Sample Resource for maintaining high-coverage whole-genome sequence data as a public resource, and the PharmCAT team for open-source pharmacogenomic annotation infrastructure.

## Footnotes

1 PharmCAT detects 2 additional *CYP2C9* *8 carriers not called by *afripharmagen*, likely reflecting a variant normalization difference at rs7900194 (left-alignment and multi-allelic decomposition by PharmCAT’s preprocessor).

